# A cross-national study of factors associated with women’s perinatal mental health and wellbeing during the COVID-19 pandemic

**DOI:** 10.1101/2020.12.03.20243519

**Authors:** Archana Basu, Hannah Hayoung Kim, Karmel W. Choi, Lily Charron, Nora Kelsall, Rebecca Basaldua, Sonia Hernandez-Diaz, Diego F. Wyszynski, Karestan C. Koenen

## Abstract

**Background:** Pregnant and postpartum women face unique challenges during the COVID-19 pandemic that may put them at elevated risk of mental health problems. However, few large-scale and no cross-national studies have been conducted to date that investigate modifiable pandemic-related behavioral or cognitive factors that may influence mental health in this vulnerable group. This international study sought to identify and measure the associations between pandemic-related information seeking, worries, and prevention behaviors on perinatal mental health during the COVID-19 pandemic.

**Methods and Findings:** An anonymous, online, cross-sectional survey of pregnant and postpartum women was conducted in 64 countries between May 26 2020 and June 13 2020. The survey, available in twelve languages, was hosted on the Pregistry platform for COVID-19 studies (https://corona.pregistry.com), and advertised predominantly in social media channels and online parenting forums. Participants completed measures on demographic characteristics, COVID-19 exposure and worries, media exposure, COVID-19 prevention behaviors, and mental health symptoms including posttraumatic stress symptoms via the IES-6, anxiety/depression via the PHQ-4, and loneliness via the UCLA-3. Of the 6,894 participants, substantial proportions of women scored at or above the cut-offs for elevated posttraumatic stress (2,979 [43%]), anxiety/depression (2,138 [31%], and loneliness (3,691 [53%]). Information seeking from any source (e.g., social media, news, talking to others) five or more times per day was associated with more than twice the odds of elevated posttraumatic stress and anxiety/depression, in adjusted models. A large majority of women (86%) reported being somewhat or very worried about COVID-19. The most commonly reported worries were related to pregnancy and delivery, including family being unable to visit after delivery (59%), the baby contracting COVID-19 (59%), lack of a support person during delivery (55%), and COVID-19 causing changes to the delivery plan (41%). Greater worries related to children (i.e. inadequate childcare, their infection risk) and missing medical appointments were associated with significantly higher odds of posttraumatic stress, anxiety/depression and loneliness. Engaging in hygiene-related COVID-19 prevention behaviors (face mask-wearing, washing hands, disinfecting surfaces) were not related to mental health symptoms or loneliness.

**Conclusions:** Clinically significant posttraumatic stress, anxiety/depression, and loneliness are highly prevalent in pregnant and postpartum women across 64 countries during the COVID-19 pandemic. Excessive information seeking and worries related to children and medical care are associated with clinically significant symptoms, whereas engaging in hygiene-related preventive measures were not. In addition to screening and monitoring mental health symptoms, reinforcing healthy information seeking, addressing worries about access to medical care and the well-being of their children, and strategies to target loneliness (e.g., online support groups) should be part of intervention efforts for perinatal women. Public and mental health interventions need to explicitly address the impact of COVID-19 on both physical and mental health in perinatal women, as prevention of viral exposure itself does not mitigate the mental health impact of the pandemic.

## Introduction

The United Nations Children’s Fund estimates that there will be 116 million births in the nine months since COVID-19 was characterized as a pandemic by the World Health Organization (WHO) on March 11 2020 [1, 2]. The unique challenges facing pregnant and postpartum women that have been studied during this time include concerns about increased severity of COVID-19 in this population, potential vertical transmission from an infected mother to her newborn, and increased risk of adverse neonatal outcomes [3, 4]. Recent studies also suggest elevated rates of mental health problems among perinatal women during the pandemic including depression [5-10], anxiety [5-13], dissociation [5], and posttraumatic stress symptoms [5, 13], as well as loneliness and isolation [14]. Research also indicates that perinatal women who have a history of psychological disorders, are experiencing higher concerns regarding their family’s health, their baby’s future and society due to the COVID-19 pandemic [15]. Meanwhile, obstetricians report that perinatal women are contacting them expressing concern about hospital visits, protection methods, their infant’s safety, anxieties due to social media and fears of contracting COVID-19 [16]. Perinatal mental health problems not only adversely impact women’s own health, but also infant outcomes [17], mother-infant bonding, and later infant physical and behavioral health outcomes [18-20]. Thus, the mental health of pregnant and postpartum women is critical for both their own well-being and that of future generations.

To date, there have been no large-scale cross-national studies of women’s perinatal mental health during the COVID-19 pandemic. Moreover, previous studies have not assessed loneliness as a mental health experience in this vulnerable group, despite the well-established importance of social connection during the perinatal period [21] and known adverse effects of loneliness on mental health, morbidity and mortality [22]. Recent work has also examined only a narrow range of behavioral and cognitive factors associated with mental health outcomes in this vulnerable group. In terms of cognitive factors such as worries and concerns, the COVID-19 pandemic has radically altered the perinatal experience as healthcare services broadly shift from in person to online and as support persons are restricted at appointments and delivery. In addition to the concerns about one’s own risk of contracting COVID-19, pregnant or postpartum women are faced with the possible effects of infection on the developing fetus or infant. Economic uncertainty coupled with increased isolation may also negatively affect maternal mental health. Finally, little is known about pregnant and postpartum women’s engagement in behaviors aimed at understanding the pandemic (e.g., information seeking) and preventing COVID-19 infection, and how such behaviors may or may not be associated with their mental health.

We conducted a survey of pregnant and postpartum women to first examine the relation of type and amount of information seeking via various channels (social media, news, discussion with others) to women’s mental health and loneliness. We then analyzed the role of pandemic-related worries in mental health and loneliness. Finally, we investigated the association between engaging in COVID-19 prevention behaviors and mental health and loneliness. The primary goal of the study was to identify modifiable behavioral and cognitive factors, which if addressed, may reduce mental health risk and improve well-being among pregnant and postpartum women.

## Methods

### Study Design and Setting

An anonymous, online, cross-sectional survey targeting pregnant and postpartum women was conducted in 64 countries between May 26 2020 and June 13 2020. Participation in this study was voluntary. All potential participants were informed about the research objectives and standards of confidentiality regarding the use of the data. The survey, hosted on the Pregistry platform for COVID-19 studies (https://corona.pregistry.com), was advertised predominantly in social media channels and online parenting forums. Advertisements and the survey were available in twelve languages (Arabic, Chinese, English, French, German, Italian, Korean, Portuguese, Russian, Spanish, Turkish, and Urdu) by human translators. sought to obtain at least 100 responses from each of the countries with the highest number of COVID-19 cases at the time of recruitment. Interested participants were invited to follow a link to take the survey. The survey collected standard demographic data and included questions that addressed topics such as COVID-19 exposure and worries, lifestyle changes, media exposure, protective factors, and mental health.

### Participants

Women who self-identified as being 18 years or older at the time of the survey and as currently pregnant or having given birth within the past 6 months were eligible to participate. The study was classified exempt by the Harvard Longwood Campus Institutional Review Board (HLC IRB) per the regulations found at 45 CFR 46.104(d)(2) on the basis that it poses no greater than minimal risk and that the recorded information cannot readily identify the subject (directly or indirectly). The total number of participants at the close of enrollment was 7,562 individuals across 64 countries.

### Measures

#### COVID-19 Assessments

##### COVID-19 Exposure

Questions assessing whether participants were tested, diagnosed, or in contact with an individual who had COVID-19 were adapted for this study based on those formulated by the US Centers for Disease Control and Prevention for the Household Pulse Survey[23].

##### COVID-19 Information Seeking

Participants were asked about the frequency (never, <1x/day, 2-4x/day, 5-8x/day, 9-16x/day, and >16x/day) of their interactions with various sources of information, including the news, social media, and interpersonal discussions about COVID-19 using a measure modified from other published studies [24, 25]. For analyses, interactions were categorized as never, <1x/day, 2-4x/day, 5+x/day.

##### COVID-19 Worries

Participants were asked to rate their overall level of worry about COVID-19 on a Likert Scale ranging from 1 for “not worried at all” to 4 for “very worried” [26]. They were then asked to endorse fifteen specific worries on a list developed for this study. Exploratory factor analyses were conducted to identify domains within the questionnaire, with oblimin rotation on a tetrachoric correlation matrix due to the binary nature of the variables. Details of the factor analyses are presented in the supplementary materials (**S2 Text**). Worries were categorized into the following domains: social (parents/grandparents unable to visit, family unable to visit, not able to have a baby shower, not able to attend a funeral), COVID-19 infection-related (participant or partner will bring infection home, family or friends will get COVID-19), child-related (no adequate childcare, other children will get COVID-19), delivery-related (partner not present during delivery, changes to delivery plan, unborn baby will get COVID-19, not able to breastfeed), economic (significantly affect economic situation/finances), and missing doctor appointments.

##### COVID-19 prevention behaviors

Participants were asked to endorse seventeen behaviors they had engaged in to protect themselves from COVID-19 from a list developed for this study based on WHO recommendations and media reports. Behaviors were classified into the following categories: hygiene-related (mask-wearing, washing hands, disinfecting surfaces), physical distancing (avoiding public places, restaurants and other people, canceling personal engagements, work or school and working at home), canceling travel (for work or pleasure), stockpiling essential resources (food or water, hand sanitizer, medication), postponing medical care, and prayer.

### Mental Health Outcomes

**Depression and Anxiety** was assessed via the **Patient Health Questionnaire-4 (PHQ-4) [27, 28]**, a four-item inventory rated on a four-point Likert-type scale. Items are drawn from the first two items of the ‘Generalized Anxiety Disorder–7 scale’ (GAD–7) and the ‘Patient Health Questionnaire-8’ (PHQ-8). The overall PHQ-4 score is a sum of the four items (0 = not at all, 1 = several days, 2 = more than half the days, 3 = nearly every day). A PHQ-4 score of >=6 is considered clinically significant.

**Posttraumatic stress symptoms** were assessed via a modified version of the **Impact of Events Scale - 6 (**IES-6). Participants were asked to report how bothered they were by each symptom from 0 (Not at all) to 4 (Extremely) over the past seven days. The symptom statements were modified to assess impact related to the COVID-19 pandemic. The scale is scored by calculating the mean of the five items used in this study, with the original IES-6 cutoff score of 1.75 yielding 0.88 sensitivity and 0.85 specificity for posttraumatic stress disorder (PTSD) diagnosis [29-31].

**Loneliness** was evaluated using the UCLA Three-Item Loneliness Scale (UCLA-3) [32]. Participants are asked how often they experience the following: feeling that they lack companionship, feeling left out, and feeling isolated from others. Questions were framed so that participants described their feelings since the start of the COVID-19 pandemic. Responses are rated as 1 (Hardly Ever), 2 (Some of the Time), or 3 (Often), with the overall score as the sum of the three responses. A score of 3 is considered low, 4-5 medium, and >= 6 high. High loneliness has been associated with poorer mental and physical health over the life course [33].

### Sociodemographic factors and potential confounders

Standard socio-demographic measures were collected including age, education (categorized as never attended school, elementary school, some high school, high school graduate or general equivalency diploma (GED), some college/university, college diploma or university degree, master’s degree, professional degree, doctoral degree), self-identified race/ethnicity (categorized as White/Caucasian, Latina/Hispanic, Asian, South Asian, Black, Middle Eastern, Native Hawaiian or Other, Pacific Islander, American Indian or Alaska Native, Other/Multiracial), employment status (healthcare worker in a hospital or clinic, worked in a nursing home, essential/key worker (as defined by the government), none of these, don’t know), medical coverage status, marital status (married, living with partner, divorced, separated, single, widowed), weeks pregnant/postpartum (first trimester 0 to <13 weeks, second trimester 13 to <28 weeks, third trimester 28+ weeks, postpartum). Participants indicated their country of residence which was classified by region for analytic purposes.

### Statistical analyses

Descriptive statistics for socio-demographic characteristics, COVID-19 exposure variables, mental health, and loneliness were calculated by perinatal stage, classified as first, second, or third trimester, or post-partum, for all participants. A series of multivariable logistic regressions were then run to test the study hypotheses. For these analyses, PHQ-4, IES-6 and UCLA-3 outcomes were specified as binary dependent variables using recommended cutoffs. The first set of models included pregnancy stage, socio-demographic variables and COVID-19 exposure as independent variables. The second set of models examined the type and amount of information seeking about COVID-19 as independent variables. The third and fourth set of models focused on COVID-19 worries and COVID-19 behaviors as independent variables. Models for information seeking, COVID-19 worries, and COVID-19 behaviors were adjusted for age, level of education, race, survey region, marital status, and pregnancy stage.

Models were estimated using the GLM function in the R statistical program, version 3.6.2. Due to the number of models run, interpretations are focused on effects that met the threshold of p< .001. Due to low levels of missingness (667 [9%]) a complete case analysis was conducted. The raw data used for the study are publicly available.

## Results

### Study population

The final analytic population consisted of 6,894 women residing in 64 countries ranging in age from 18 to 46 years. Due to the convenience sampling design, information on response rate is not available.

Demographic characteristics are reported in **Table 1**. The mean age of the respondents was 31.3 (SD: 4.8) years. The largest proportion of respondents reported being from South or Latin America (29%), with Europe a close second (24%), followed by North America (18%), Asia & Pacific (17%), Africa (10%), and the Middle East (2%). The majority of women identified their race as White (46%) or Latin/Hispanic (21%) with fewer women identifying as Asian (13%) or Black (8%) and the remaining women identifying as South Asian (12%), Middle Eastern (1%), Native/Indigenous (<0%), more than one race (4%) or other/not specified (3%) with a small number of women missing race information but who answered all other questions (1%). The majority of women reported being married (65%), being at least a college graduate (73%), not being an essential or healthcare worker (75%), and having health care coverage (71%). With regard to COVID-19 exposure, the vast majority of women reported not having been tested for COVID-19 (89%), not having been in contact with an individual who has or had COVID-19 (78%), never having been diagnosed with COVID-19 (98%), and not being an essential or healthcare worker (75%).

**Table 1.** Descriptive statistics for survey respondents in the analytic sample, overall and by pregnancy status.

| Variable | Gestational weeks at participation |  |  |  |  |
| --- | --- | --- | --- | --- | --- |
|  | 0 to <13 weeks<br>(n=1,276) | 13 to <28 weeks<br>(n=2,443) | 28+ weeks<br>(n=1,993) | Postpartum<br>(n=1,182) | Overall<br>(n=6,894) |
| **Age in years** |  |  |  |  |  |
| Mean (SD) | 30.2 (4.92) | 31.4 (4.71) | 31.7 (4.77) | 31.7 (4.94) | 31.3 (4.84) |
| Median [Min, Max] | 30.0 [18.0, 44.0] | 31.0 [18.0, 44.0] | 32.0 [18.0, 46.0] | 32.0 [18.0, 46.0] | 32.0 [18.0, 46.0] |
| **Region** |  |  |  |  |  |
| Africa | 137 (10.7%) | 284 (11.6%) | 204 (10.2%) | 90 (7.6%) | 715 (10.4%) |
| Asia & Pacific | 333 (26.1%) | 306 (12.5%) | 206 (10.3%) | 320 (27.1%) | 1,165 (16.9%) |
| Europe | 186 (14.6%) | 626 (25.6%) | 517 (25.9%) | 304 (25.7%) | 1,633 (23.7%) |
| Middle East | 18 (1.4%) | 48 (2.0%) | 29 (1.5%) | 16 (1.4%) | 111 (1.6%) |
| North America | 194 (15.2%) | 368 (15.1%) | 405 (20.3%) | 278 (23.5%) | 1,245 (18.1%) |
| South/Latin America | 408 (32.0%) | 811 (33.2%) | 632 (31.7%) | 174 (14.7%) | 2,025 (29.4%) |
| **Race/ethnicity** |  |  |  |  |  |
| White | 464 (36.4%) | 1,134 (46.4%) | 995 (49.9%) | 582 (49.2%) | 3,175 (46.1%) |
| Latin/Hispanic | 285 (22.3%) | 565 (23.1%) | 438 (22.0%) | 131 (11.1%) | 1,419 (20.6%) |
| Asian | 261 (20.5%) | 223 (9.1%) | 152 (7.6%) | 262 (22.2%) | 898 (13.0%) |
| Black | 123 (9.6%) | 215 (8.8%) | 169 (8.5%) | 74 (6.3%) | 581 (8.4%) |
| South Asian | 28 (2.2%) | 42 (1.7%) | 37 (1.9%) | 21 (1.8%) | 128 (1.9%) |
| Middle Eastern | 11 (0.9%) | 34 (1.4%) | 27 (1.4%) | 17 (1.4%) | 89 (1.3%) |
| Native/Indigenous | 7 (0.5%) | 7 (0.3%) | 6 (0.3%) | 4 (0.3%) | 24 (0.3%) |
| More than 1 race/ethnicity | 45 (3.5%) | 108 (4.4%) | 95 (4.8%) | 45 (3.8%) | 293 (4.3%) |
| Other | 43 (3.4%) | 83 (3.4%) | 58 (2.9%) | 33 (2.8%) | 217 (3.1%) |
| Not reported | 9 (0.7%) | 32 (1.3%) | 16 (0.8%) | 13 (1.1%) | 70 (1.0%) |
| **Marital status** |  |  |  |  |  |
| Married | 811 (63.6%) | 1,518 (62.1%) | 1,292 (64.8%) | 852 (72.1%) | 4,473 (64.9%) |
| Living with partner | 355 (27.8%) | 725 (29.7%) | 507 (25.4%) | 249 (21.1%) | 1,836 (26.6%) |
| Other | 110 (8.6%) | 200 (8.2%) | 194 (9.7%) | 81 (6.9%) | 585 (8.5%) |
| **Level of education** |  |  |  |  |  |
| High school graduate or less | 164 (12.9%) | 335 (13.7%) | 281 (14.1%) | 142 (12.0%) | 922 (13.4%) |
| Some college | 205 (16.1%) | 363 (14.9%) | 280 (14.0%) | 122 (10.3%) | 970 (14.1%) |
| College graduate | 462 (36.2%) | 871 (35.7%) | 720 (36.1%) | 511 (43.2%) | 2,564 (37.2%) |
| Graduate school or more | 445 (34.9%) | 874 (35.8%) | 712 (35.7%) | 407 (34.4%) | 2,438 (35.4%) |
| **Employment status** |  |  |  |  |  |
| Worked in healthcare or nursing home | 144 (11.3%) | 301 (12.3%) | 225 (11.3%) | 120 (10.2%) | 790 (11.5%) |
| Essential worker | 189 (14.8%) | 378 (15.5%) | 273 (13.7%) | 113 (9.6%) | 953 (13.8%) |
| Other | 943 (73.9%) | 1,764 (72.2%) | 1,495 (75.0%) | 949 (80.3%) | 5,151 (74.7%) |
| **Medical coverage status** |  |  |  |  |  |
| Yes | 885 (69.4%) | 1,699 (69.5%) | 1,442 (72.4%) | 873 (73.9%) | 4,899 (71.1%) |
| **Tested for COVID-19** |  |  |  |  |  |
| Negative, I did not have the virus | 130 (10.2%) | 139 (5.7%) | 134 (6.7%) | 238 (20.1%) | 641 (9.3%) |
| No, I have not been tested | 1,114 (87.3%) | 2,260 (92.5%) | 1,824 (91.5%) | 913 (77.2%) | 6,111 (88.6%) |
| Positive, I had the virus | 14 (1.1%) | 23 (0.9%) | 17 (0.9%) | 8 (0.7%) | 62 (0.9%) |
| Yes, but I do not know the result yet or the result was inconclusive | 18 (1.4%) | 21 (0.9%) | 18 (0.9%) | 23 (1.9%) | 80 (1.2%) |
| **In contact with an individual who has COVID-19** |  |  |  |  |  |
| No | 972 (76.2%) | 1,887 (77.2%) | 1,564 (78.5%) | 943 (79.8%) | 5,366 (77.8%) |
| Maybe | 198 (15.5%) | 371 (15.2%) | 287 (14.4%) | 162 (13.7%) | 1,018 (14.8%) |
| Yes | 106 (8.3%) | 185 (7.6%) | 142 (7.1%) | 77 (6.5%) | 510 (7.4%) |
| **Diagnosed with COVID-19** |  |  |  |  |  |
| No | 1,246 (97.6%) | 2,394 (98.0%) | 1,971 (98.9%) | 1,166 (98.6%) | 6,777 (98.3%) |
| Yes, and I still have it | 12 (0.9%) | 12 (0.5%) | 5 (0.3%) | 3 (0.3%) | 32 (0.5%) |
| Yes, but I recovered | 18 (1.4%) | 37 (1.5%) | 17 (0.9%) | 13 (1.1%) | 85 (1.2%) |
| **IES-6 score** |  |  |  |  |  |
| <1.75 | 751 (58.9%) | 1,367 (56.0%) | 1,149 (57.7%) | 648 (54.8%) | 3,915 (56.8%) |
| >=1.75 | 525 (41.1%) | 1,076 (44.0%) | 844 (42.3%) | 534 (45.2%) | 2,979 (43.2%) |
| **PHQ4 score** |  |  |  |  |  |
| <6 | 850 (66.6%) | 1,702 (69.7%) | 1,385 (69.5%) | 819 (69.3%) | 4,756 (69.0%) |
| >=6 | 426 (33.4%) | 741 (30.3%) | 608 (30.5%) | 363 (30.7%) | 2,138 (31.0%) |
| **UCLA-3 score** |  |  |  |  |  |
| <6 | 646 (50.6%) | 1,146 (46.9%) | 888 (44.6%) | 523 (44.2%) | 3,203 (46.5%) |
| >=6 | 630 (49.4%) | 1,297 (53.1%) | 1,105 (55.4%) | 659 (55.8%) | 3,691 (53.5%) |

Abbreviations: SD, standard deviation

### Mental health and loneliness

Substantial proportions of women scored at or above the risk cut-offs for IES-6 (2,979 [43%]), PHQ-4 (2,138 [31%]), and UCLA-3 (3,691 [54%]). **Fig 2** presents the proportions of women in our sample at or above clinical risk cut-offs for elevated PTSD, anxiety and depression compared with other studies of pregnant and postpartum women from meta-analysis and during COVID-19 as well as the US population modified from that presented by Lebel et al [7].

**Fig 1.**
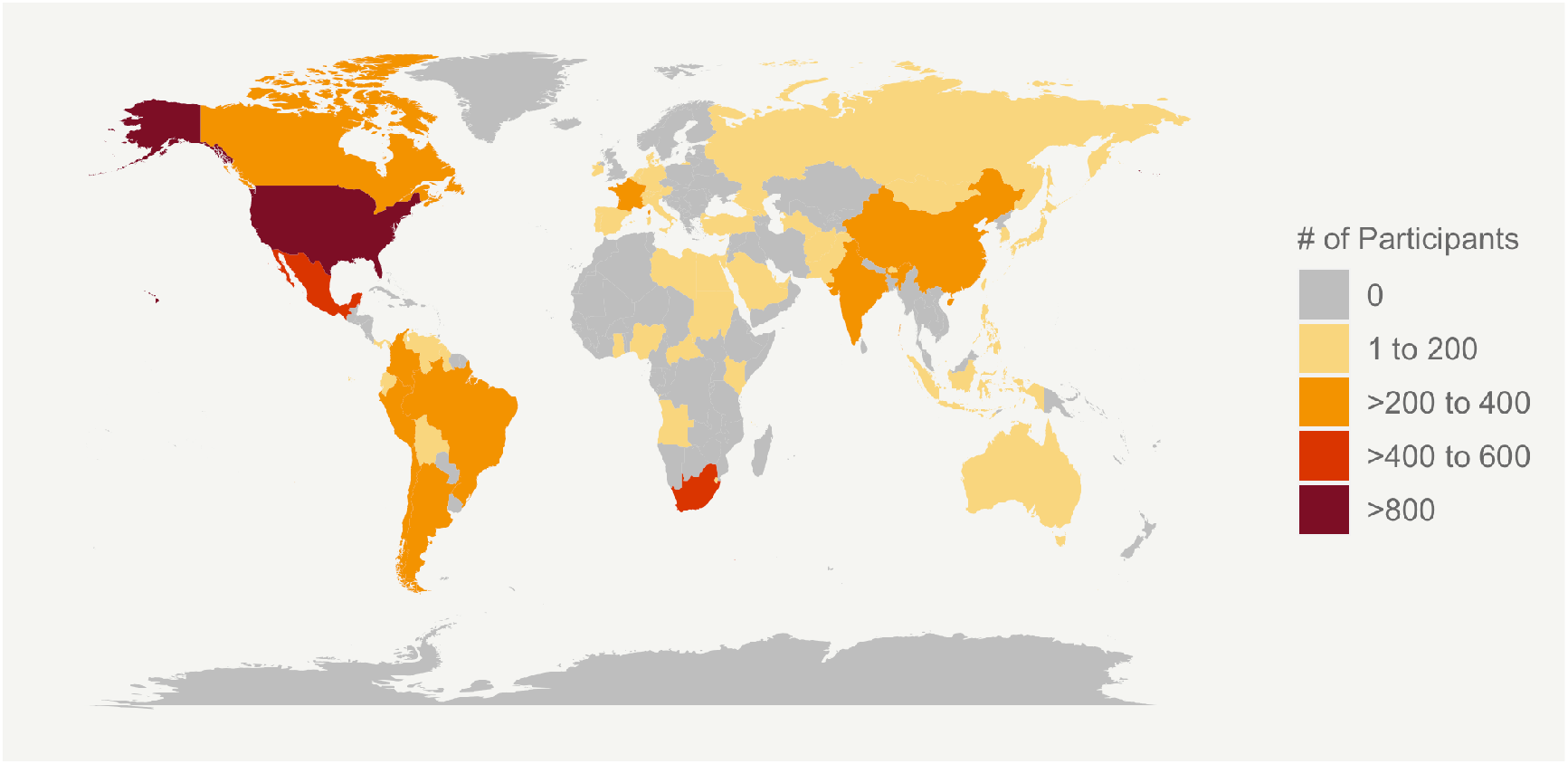
Geographic distribution of study participants by country.

**Fig 2.**
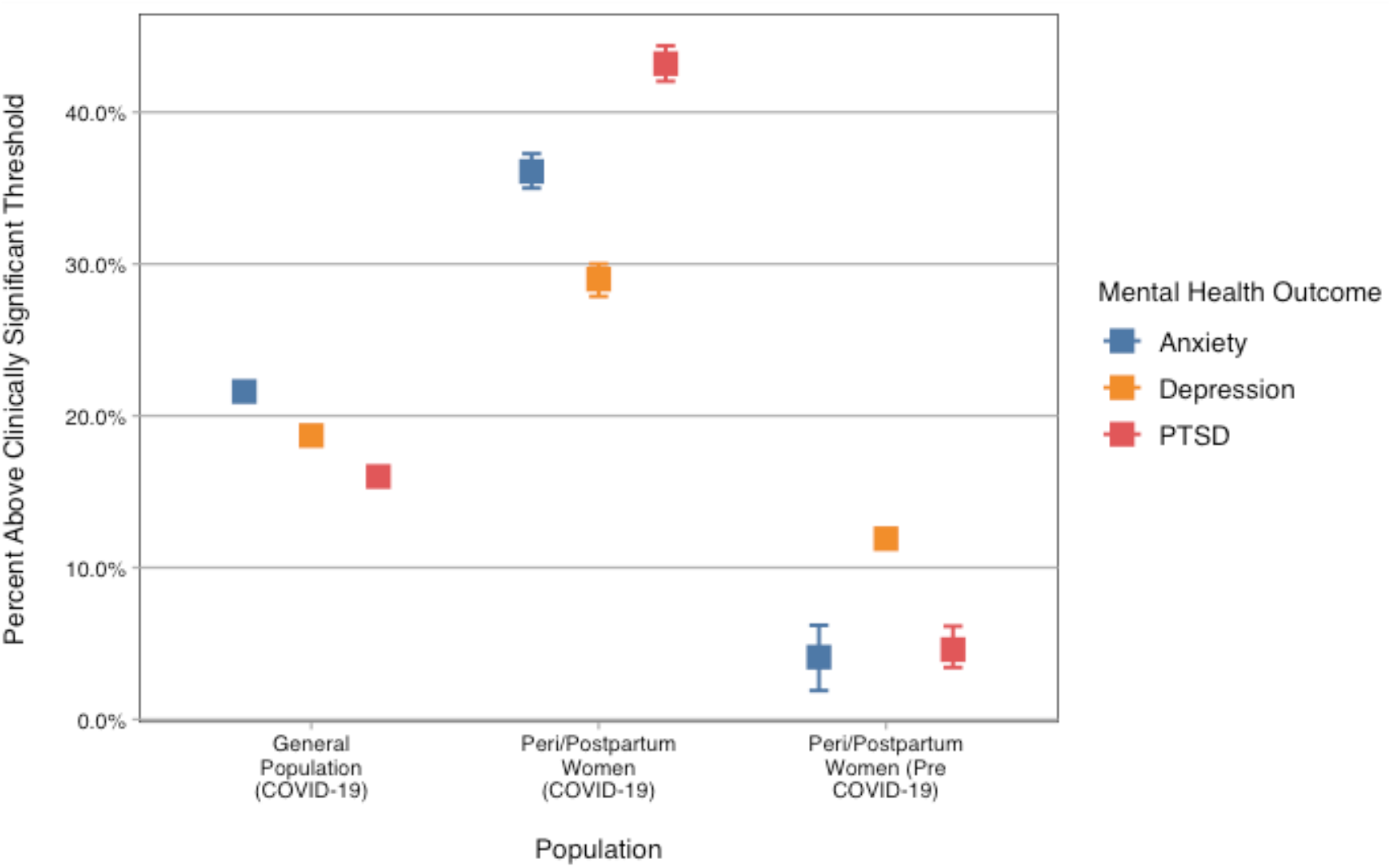
Prevalence of clinically significant anxiety, depression and posttraumatic stress in the general population during COVID1-9 and in perinatal women before COVID-19 and during COVID-19 in our survey [18, 19, 34-36].

Results for multivariable models of socio-demographic and COVID-19 exposure variables in relation to IES-6, PHQ4 and UCLA-3 are presented in the supplementary materials (see **Table A in S1 Text**). In fully adjusted models, only age was consistently associated with reduced risk of clinically significant outcomes across measures at p<.001. Only “maybe” having been in contact with someone with COVID-19 compared to “never” was consistently associated with increased risk of clinically significant outcomes across measures at p<.001.

### Information Seeking and Mental Health

**Table 2** presents the results of multivariable logistic regression models for specific types of information seeking in relation to IES-6, PHQ4 and UCLA-3.

**Table 2.**
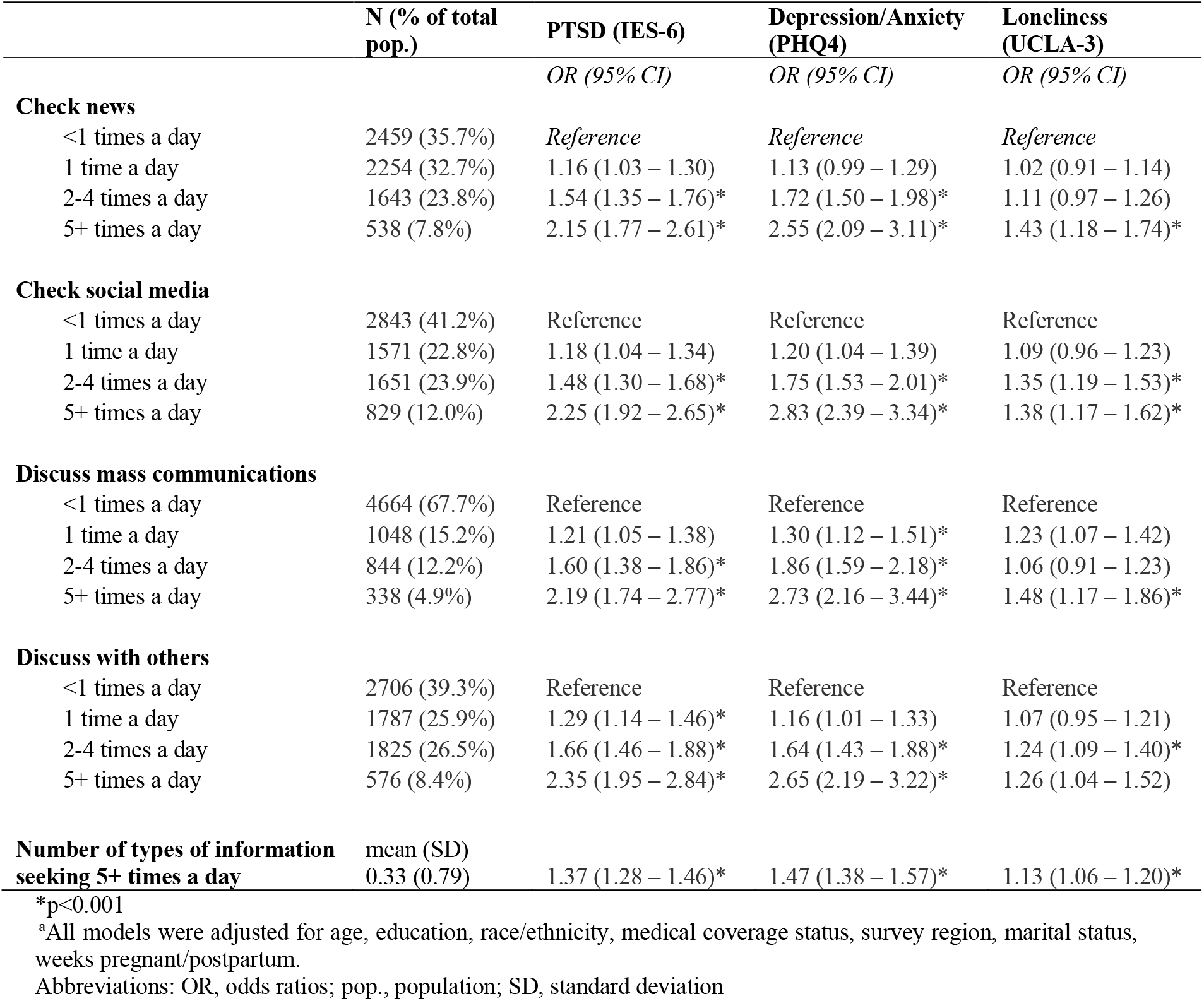
Model results for PTSD, depression/anxiety, and loneliness in relation to frequency of different types of information seeking^a^.

For all types of information seeking, there was a dose-response relation between information seeking frequency and meeting the clinical threshold for both the IES-6 and PHQ-4, in fully adjusted models. In fact, participants engaging in any type of information seeking five or more times per day had more than twice the odds of meeting the clinical threshold for the IES-6 and PHQ-4 (**Table 2**) compared to those who did not engage in that type of information seeking. There was also a dose-response relation between number of types of information seeking 5+ times a day and meeting the clinical threshold on the IES-6 and PHQ-4, with individuals seeking information via multiple sources showing more than twice the odds of distress.

Information seeking was less strongly and consistently related to meeting the risk threshold on the UCLA-3. Checking news, social media, and mass communications increased the odds of loneliness between 38 to 48% (**Table 2**) for women doing so 5+ times a day compared to women who engaged in information seeking less than once a day. Discussing with others was only associated with increased loneliness when done 2-4x per day.

### Worries and Mental Health

The prevalence of reported COVID-19 related worries is presented in **Fig 3** and **Table B in S1 Text**. A high percentage of respondents (86%) reported being somewhat or very worried about COVID-19, with many of the most commonly reported worries related to pregnancy and delivery including: family being unable to visit after delivery (59%), the baby contracting COVID-19 (59%), lack of a support person during delivery (55%), and COVID-19 causing changes to the delivery plan (41%). Being very worried about COVID-19, compared to not having any worries, was strongly and significantly associated with meeting the clinical cut-off both on the IES-6 (OR=4.75, 95% CI: 3.34, 6.87) and the PHQ-4 (OR=1.51, 95% CI: 1.09, 2.13) but was not associated with the UCLA-3 (OR=1.25, 95% CI: 0.90, 1.72), in fully adjusted models.

**Fig 3.**
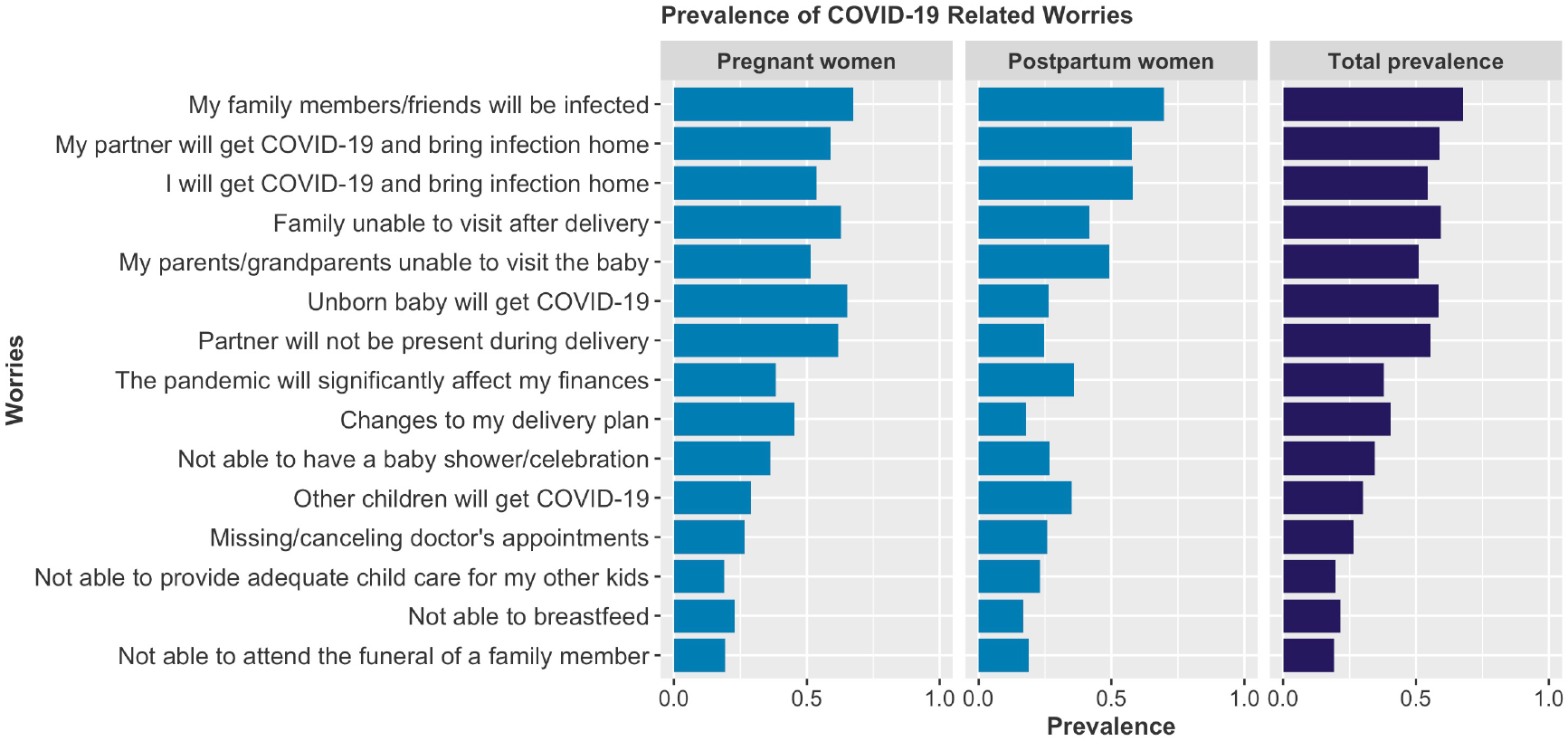
Prevalence of COVID-19 related worries for pregnant and postpartum women.

Using domains obtained from the factor analysis (see **S2 Text**), **Table 3** presents the association between mental health outcomes and the six COVID-19 worry categories. Child-related worries and medical care worries were each associated with significantly higher odds of exceeding the clinical threshold on the IES-6, PHQ4 and UCLA-3. Social worries were only associated with higher odds of loneliness. Economic worries were associated with increased odds of meeting the clinical threshold for IES-6 and PHQ4; infection and delivery worries were only significantly associated with increased odds of meeting the clinical threshold on the IES-6. The prevalence of PTSD, depression/anxiety, and loneliness consistently increased for every additional worry category endorsed (**Fig 4**).

**Table 3.**
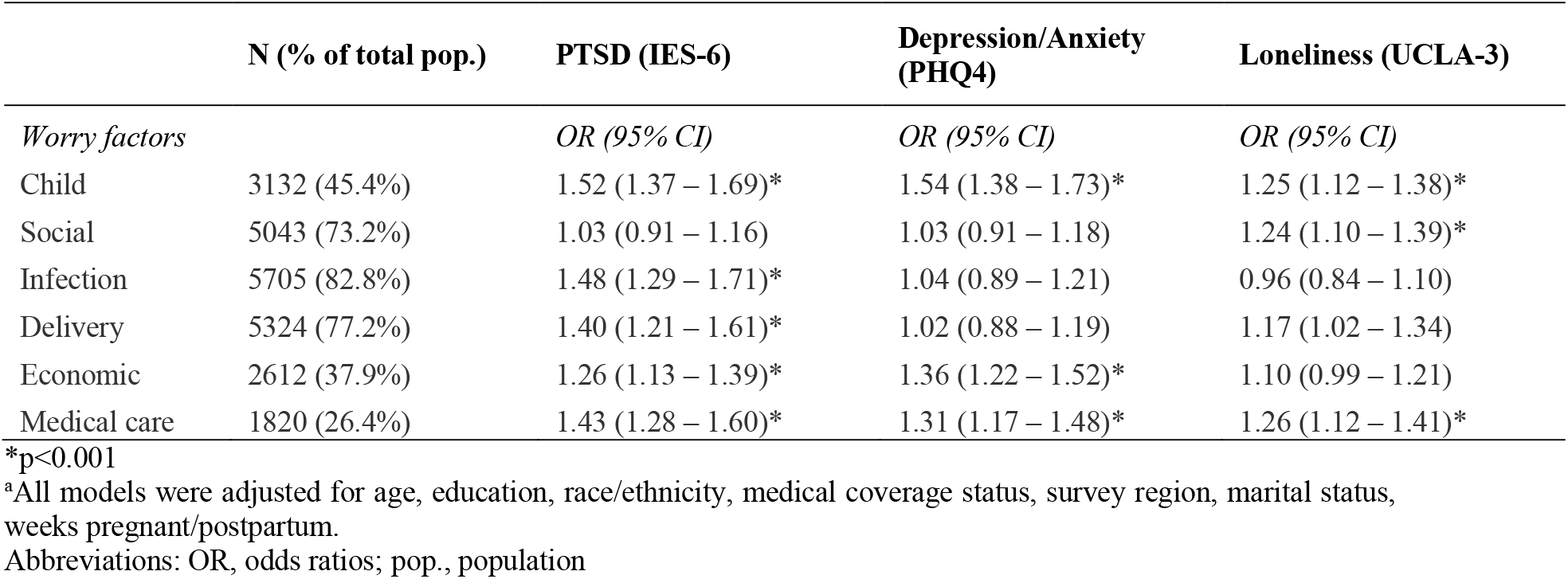
Results of multivariable logistic regression models for PTSD, depression/anxiety, and loneliness in relation to the four COVID-19 worry factors: child, social, infection, delivery as well as economic worries and missing doctors’ appointments^a^.

**Fig 4.**
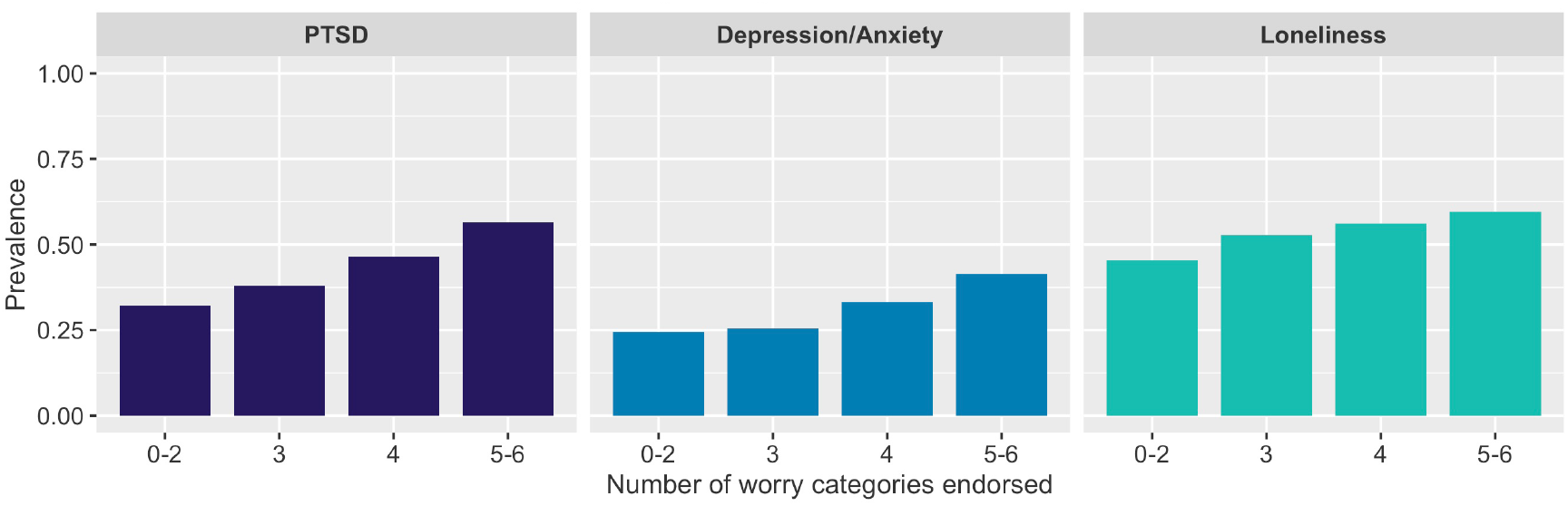
Prevalence of PTSD, depression/anxiety, and loneliness by the number of COVID-19-related worry categories endorsed by pregnant and postpartum women.

### Behaviors and Mental Health

The prevalence of reported engagement in COVID-19 prevention related behaviors is presented in **Fig 5** and further details are provided in **Table C in S1 Text**. The majority of women engaged in COVID-19 prevention behaviors recommended by public health experts: 93% washed/sanitized hands several times per day, 85% wore a face mask, 83% avoided crowds, 70% avoided eating in restaurants, 67% avoided contact with high-risk people, 64% disinfected surfaces, and 55% canceled or postponed activities. This pattern of engagement was largely consistent across pregnancy stage. Results of logistic regression models for IES-6, PHQ-4 and UCLA-3 in relation to types of COVID-19 prevention behaviors are presented in **Table 4**.

**Table 4.**
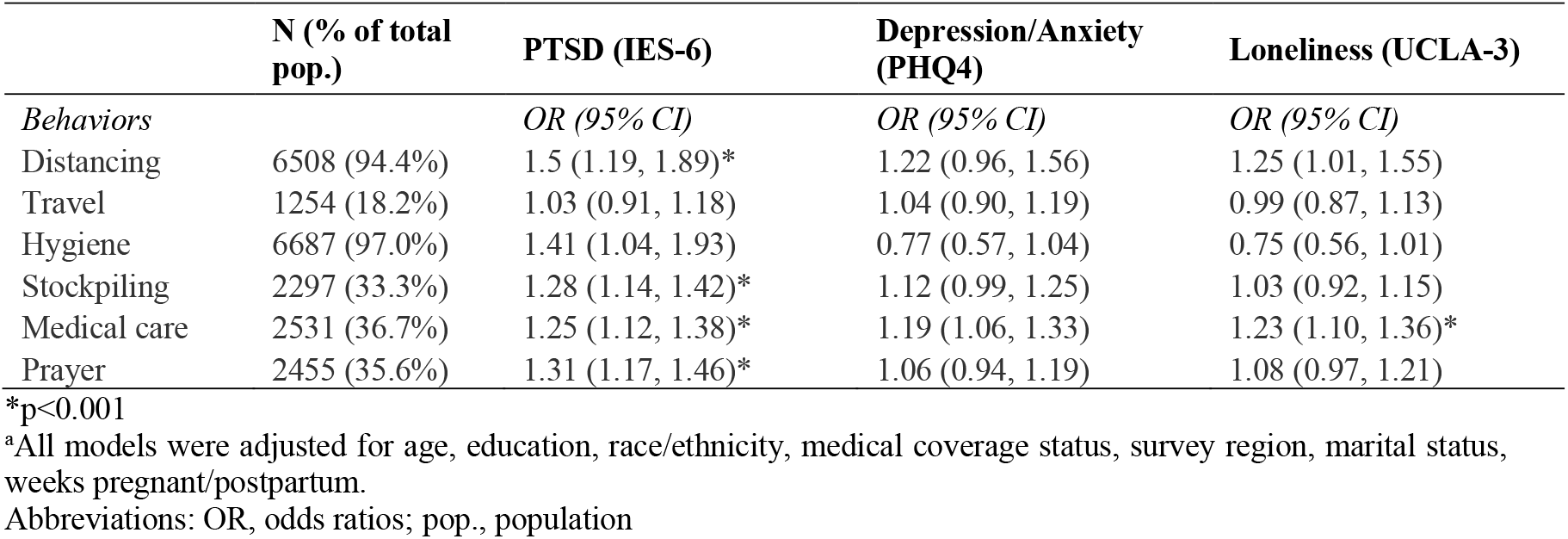
Results of logistic regression models for PTSD, depression/anxiety, and loneliness in relation to the six COVID-19 behavior groups: distancing, travel, hygiene, stockpiling essential resources, medical care seeking, and prayer^a^.

|  | N (% of total pop.) | PTSD (IES-6) | Depression/Anxiety (PHQ4) | Loneliness (UCLA-3) |
| --- | --- | --- | --- | --- |
| *Behaviors* |  | *OR (95% CI)* | *OR (95% CI)* | *OR (95% CI)* |
| Distancing | 6508 (94.4%) | 1.5 (1.19, 1.89)* | 1.22 (0.96, 1.56) | 1.25 (1.01, 1.55) |
| Travel | 1254 (18.2%) | 1.03 (0.91, 1.18) | 1.04 (0.90, 1.19) | 0.99 (0.87, 1.13) |
| Hygiene | 6687 (97.0%) | 1.41 (1.04, 1.93) | 0.77 (0.57, 1.04) | 0.75 (0.56, 1.01) |
| Stockpiling | 2297 (33.3%) | 1.28 (1.14, 1.42)* | 1.12 (0.99, 1.25) | 1.03 (0.92, 1.15) |
| Medical care | 2531 (36.7%) | 1.25 (1.12, 1.38)* | 1.19 (1.06, 1.33) | 1.23 (1.10, 1.36)* |
| Prayer | 2455 (35.6%) | 1.31 (1.17, 1.46)* | 1.06 (0.94, 1.19) | 1.08 (0.97, 1.21) |

\*p<0.001
<sup>a</sup>All models were adjusted for age, education, race/ethnicity, medical coverage status, survey region, marital status, weeks pregnant/postpartum.
Abbreviations: OR, odds ratios; pop., population

**Fig 5.**
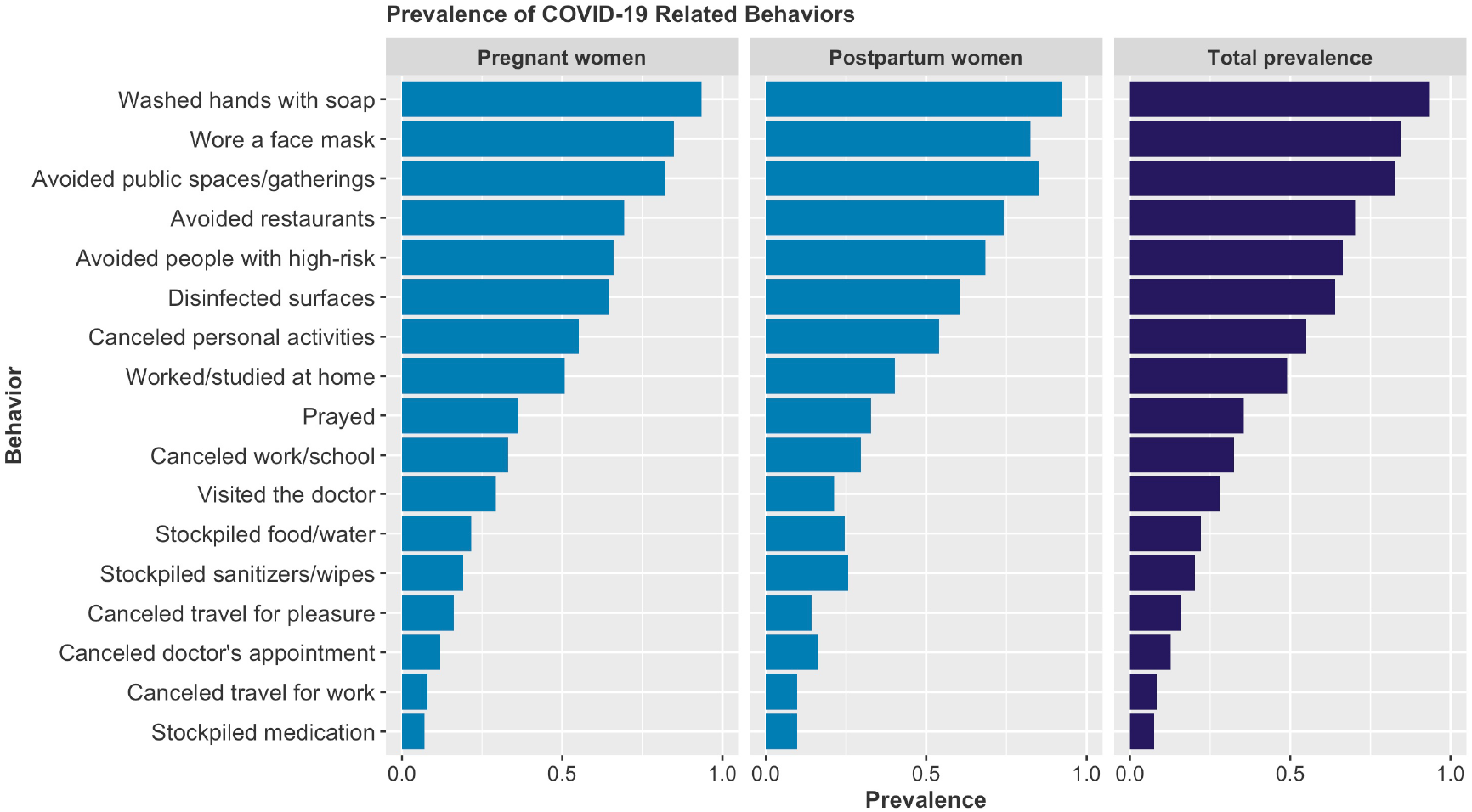
Prevalence of COVID-19 prevention related behaviors for pregnant and postpartum women.

Hygiene and travel behaviors were not related to clinically elevated IES-6, PHQ-4, or UCLA-3. However, distancing and stockpiling behaviors as well as canceling doctor’s appointments and prayer were associated with increased odds of meeting the clinical threshold on the IES-6. None of the COVID-19 related prevention behaviors were associated with increased odds of meeting the PHQ-4 clinical threshold. Only canceling doctor’s appointments was associated with increased loneliness.

## Discussion

In this large-scale anonymous, online, international cross-sectional survey of pregnant and postpartum women from 64 countries during the COVID-19 pandemic, the prevalence of clinically elevated mental health symptoms were high, with 43%, 31%, and 54% of women exceeding clinical risk thresholds for PTSD, depression/anxiety, and loneliness, respectively. Excessive pandemic-related information seeking was the strongest correlate of adverse mental health outcomes. Across any modality or type of media, information seeking five or more times per day, was associated with a more than two-fold increased risk of clinical elevations in posttraumatic stress and depression/anxiety. Moreover, there was no evidence that discussion about COVID-19, whether with another person or in social media, reduced loneliness. The vast majority of women in our study reported feeling somewhat to very worried about COVID-19, with large proportions reporting pregnancy or birth specific worries such as lack of social support around delivery. Child-related worries (inadequate childcare, other children will get COVID-19) and worries about missing doctor appointments were independently associated with increased odds of clinical elevations in all outcomes. Most women reported following public health science supported COVID-19 hygiene behaviors (e.g., face mask wearing, washing hands). Such behaviors were not associated with adverse mental health symptoms or loneliness.

Prevalence rates of psychiatric distress in our study exceed non-pandemic perinatal studies of prenatal and postpartum depression and anxiety [18, 19] and most general population estimates during pandemics [37]. Findings are especially notable as a very small proportion of women report COVID-19 infection and most women (> 75%) also reported that they were not exposed to anyone with a known infection. Recent commentaries highlight that women may be particularly vulnerable to the impact of the pandemic’s effects and public health measures taken to reduce its effects, e.g., may be more likely to lose employment due to child care demands given school closures [38]. These considerations may be particularly salient for pregnant and postpartum women, for whom pandemic associated worries and the physical distancing required for infection control may exacerbate mental health problems and feelings of loneliness. Given the potential implications for maternal and child health, acknowledging the pandemic’s unique impact on pregnant and postpartum women is central to tailoring and scaling public and mental health intervention efforts to address their particular needs.

Our findings on information seeking are consistent with others that have found high levels of pandemic-related information-seeking, whether social media or traditional news sources, is associated with increased prevalence of anxiety and depression [24, 25]. Our results extend these findings to perinatal women and expand the modalities examined. Strikingly, we found that even discussing COVID-19 *with another person* five or more times a day was associated with a two-fold increase in posttraumatic stress and depression/anxiety. Discussing COVID-19, whether with another person or in mass communications (e.g. Facebook, Twitter) was not associated with reduced loneliness. In fact, there was evidence of higher loneliness in persons who engaged in such discussions frequently, suggesting that interpersonal engagement with others, while generally beneficial for mental health, may not be protective if strongly focused on the pandemic itself.

Degree of worry about COVID-19 was associated with clinically elevated symptoms of posttraumatic stress and depression/anxiety but not loneliness. With regard to specific worries, child-related worries and missing medical appointments were consistently associated elevations in posttraumatic stress, depression/anxiety and loneliness. At the time of this study, we identified only one other study of pregnant or postpartum women that reported on specific worries [39]. Our findings, based on a data-driven approach to identify worry domains, were consistent with those reported by Corbett et al., who found that a majority of women had worries about the health of their relatives, children, and the unborn baby. Additionally, our results show that substantial proportions of women also have worries related to their delivery plan, with specific concerns about not being allowed a support person during delivery. Providing women with up-to-date, accurate information on how to prevent infection of themselves and their children, helping women understand the steps healthcare systems are taking to prevent infections of patients, especially newborns, and providing consistent prenatal and postpartum care may help support pregnant women’s mental health during this time.

With respect to behaviors to safeguard against the pandemic, most women reported engaging in prevention efforts recommended by public health experts i.e. hand hygiene, wearing a face mask, avoiding crowds and public places such as restaurants, avoiding contact with high-risk individuals, and disinfecting surfaces. Most notably, hygiene-promoting behaviors were not associated with clinically significant mental disorder symptoms or loneliness. While hygiene promotion behaviors are central to containing the viral spread and mitigating risk, engaging in such behaviors (or not) are unrelated to mental health. This has important implications for clinical research and intervention efforts as it suggests that different interventions may be needed to address the infectious disease aspects of the pandemic versus the mental health and emotional well-being aspects. Associations between other pandemic-related prevention behaviors and mental health and loneliness were inconsistent.

Our study findings should be interpreted in light of some limitations. The cross-sectional nature of the study prevents any causal attributions between the factors we examined. For instance, distancing behaviors and postponing medical care to prevent the contraction of COVID-19 (but not changes to travel plans and hygiene behaviors) were associated with clinically significant posttraumatic stress. Women who experience elevated posttraumatic stress symptoms may be more likely to isolate themselves; social avoidance is part of the syndrome. Social distancing efforts may also perpetuate or exacerbate posttraumatic stress [40]. Additionally, our study findings are based on a convenience sample and thus not-representative of any country or region. Women in our study are likely those more active on social media platforms, which was our primary modality of recruitment. Accordingly, our study likely also does not represent a comprehensive range of concerns and mental health needs of pregnant or postpartum women, such as those with limited internet access. However, given the large sample size, the availability of the survey in multiple languages, and the paucity of perinatal mental health data for women during the COVID-19 pandemic, our study provides initial information for future research and intervention efforts.

## Conclusion

Pregnant and postpartum women are reporting clinically significant depression, anxiety, and posttraumatic stress, which are higher than most available general population estimates of psychiatric distress during the pandemic. Such high levels of mental health problems have potential implications for women, and fetal and child health and development. Thus, efforts to screen, monitor, and target a range of mental health symptoms, and other aspects of emotional well-being, such as feeling of loneliness and worries, and coping behaviors (e.g., appropriate information seeking) should be considered. Reinforcing the importance of appropriate information seeking should be a key consideration in intervention efforts. Future studies may also consider examining women’s specific concerns and worries related to child well-being and concerns about access to medical care. Finally, public and mental health interventions need to explicitly address both the viral disease risks as well as the mental health risks associated with the pandemic, as prevention of viral exposure itself may not mitigate the pandemic’s mental health impact.

## Data Availability

Data is available upon request. All data requests should be directed to Dr. Karestan Koenen via email

