## Supplemental Materials for "A cross-national study of factors associated with women’s perinatal mental health and wellbeing during the COVID-19 pandemic"

**Supplementary Materials (S1)**

**Table A. Results of logistic regression models for PTSD, depression/anxiety, and loneliness in relation to socio-demographic characteristics and COVID-19 exposure.**

| | **PTSD (IES-6)** | | **Depression/Anxiety (PHQ4)** | | **Loneliness (UCLA-3)** | |
|---|---|---|---|---|---|---|
| *Predictors* | *OR (95% CI)* | *P value* | *OR (95% CI)* | *P value* | *OR (95% CI)* | *P value* |
| (Intercept) | 0.72 (0.47 – 1.09) | 0.124 | 1.43 (0.91 – 2.23) | 0.121 | 1.64 (1.09 – 2.49) | 0.019 |
| Age in years | 0.98 (0.97 – 0.99) | <0.001 | 0.96 (0.95 – 0.97) | <0.001 | 0.98 (0.97 – 0.99) | 0.001 |
| Education | | | | | | |
| High school graduate or less | *Reference* | | *Reference* | | *Reference* | |
| Some college | 1.10 (0.91 – 1.33) | 0.307 | 0.97 (0.80 – 1.18) | 0.779 | 0.92 (0.76 – 1.11) | 0.386 |
| College graduate | 1.19 (1.02 – 1.40) | 0.032 | 0.87 (0.73 – 1.02) | 0.091 | 1.00 (0.85 – 1.17) | 0.992 |
| Graduate school or more | 1.15 (0.97 – 1.35) | 0.101 | 0.80 (0.68 – 0.95) | 0.012 | 0.91 (0.78 – 1.07) | 0.272 |
| Race/ethnicity | | | | | | |
| White | *Reference* | | *Reference* | | *Reference* | |
| Latin/Hispanic | 1.09 (0.90 – 1.32) | 0.363 | 0.82 (0.67 – 1.00) | 0.047 | 0.74 (0.61 – 0.89) | 0.002 |
| Asian | 1.74 (1.36 – 2.24) | <0.001 | 1.27 (0.98 – 1.67) | 0.076 | 0.84 (0.66 – 1.06) | 0.144 |
| Black | 1.34 (1.07 – 1.68) | 0.012 | 1.03 (0.81 – 1.31) | 0.798 | 0.79 (0.63 – 0.99) | 0.040 |
| South Asian | 1.72 (1.14 – 2.57) | 0.009 | 0.90 (0.56 – 1.41) | 0.649 | 1.02 (0.69 – 1.53) | 0.917 |
| Middle Eastern | 1.14 (0.68 – 1.87) | 0.620 | 1.00 (0.58 – 1.69) | 0.990 | 0.45 (0.27 – 0.73) | 0.002 |
| Native/Indigenous | 1.53 (0.67 – 3.50) | 0.312 | 0.97 (0.40 – 2.25) | 0.947 | 0.35 (0.14 – 0.81) | 0.017 |
| More than 1 | 1.04 (0.80 – 1.33) | 0.783 | 0.94 (0.71 – 1.22) | 0.629 | 0.78 (0.61 – 1.00) | 0.054 |
| Other | 1.00 (0.74 – 1.35) | 0.991 | 0.78 (0.57 – 1.08) | 0.138 | 0.67 (0.50 – 0.90) | 0.008 |
| Missing indicator | 1.78 (1.07 – 3.00) | 0.027 | 0.97 (0.56 – 1.65) | 0.918 | 1.16 (0.70 – 1.96) | 0.560 |
| Medical coverage status | | | | | | |
| No | *Reference* | | *Reference* | | *Reference* | |
| Yes | 0.92 (0.82 – 1.03) | 0.149 | 0.70 (0.63 – 0.79) | <0.001 | 0.94 (0.84 – 1.05) | 0.258 |
| Region | | | | | | |
| Asia & Pacific | *Reference* | | *Reference* | | *Reference* | |
| Africa | 1.91 (1.43 – 2.56) | <0.001 | 1.54 (1.13 – 2.10) | 0.006 | 1.36 (1.03 – 1.81) | 0.031 |
| Europe | 1.52 (1.18 – 1.95) | 0.001 | 0.92 (0.70 – 1.21) | 0.566 | 1.32 (1.04 – 1.68) | 0.023 |
| Middle East | 3.65 (2.20 – 6.15) | <0.001 | 2.15 (1.29 – 3.59) | 0.003 | 1.45 (0.89 – 2.38) | 0.139 |
| North America | 1.60 (1.25 – 2.07) | <0.001 | 1.31 (1.00 – 1.72) | 0.055 | 1.66 (1.31 – 2.12) | <0.001 |
| South/Latin America | 1.45 (1.10 – 1.90) | 0.008 | 1.81 (1.36 – 2.42) | <0.001 | 1.12 (0.86 – 1.45) | 0.413 |
| Marital status | | | | | | |
| Married | *Reference* | | *Reference* | | *Reference* | |
| Living with partner | 0.93 (0.83 – 1.05) | 0.226 | 1.04 (0.91 – 1.18) | 0.575 | 1.01 (0.90 – 1.14) | 0.839 |
| Other | 1.16 (0.97 – 1.40) | 0.109 | 1.59 (1.31 – 1.91) | <0.001 | 1.80 (1.49 – 2.18) | <0.001 |
| Weeks pregnant/postpartum | | | | | | |
| 0 to <13 weeks | *Reference* | | *Reference* | | *Reference* | |

| | | | | | | |
|---|---|---|---|---|---|---|
| 13 to <28 weeks | 1.16 (1.01 – 1.33) | 0.041 | 0.93 (0.80 – 1.08) | 0.348 | 1.12 (0.98 – 1.29) | 0.106 |
| 28+ weeks | 1.10 (0.95 – 1.27) | 0.215 | 0.97 (0.83 – 1.13) | 0.669 | 1.21 (1.04 – 1.40) | 0.011 |
| Postpartum | 1.27 (1.08 – 1.50) | 0.004 | 1.06 (0.89 – 1.26) | 0.539 | 1.28 (1.09 – 1.51) | 0.003 |
| Tested for COVID-19 | | | | | | |
| No, I have not been tested | *Reference* | | *Reference* | | *Reference* | |
| Negative, I did not have the virus | 0.78 (0.65 – 0.93) | 0.005 | 1.02 (0.85 – 1.23) | 0.814 | 0.81 (0.68 – 0.96) | 0.014 |
| Positive, I had the virus | 0.72 (0.40 – 1.27) | 0.259 | 1.39 (0.77 – 2.50) | 0.265 | 0.92 (0.52 – 1.64) | 0.776 |
| Yes, but I do not know the results yet or the result was inconclusive | 1.33 (0.84 – 2.13) | 0.221 | 0.98 (0.59 – 1.59) | 0.934 | 0.70 (0.44 – 1.10) | 0.123 |
| In contact with someone who has/had COVID-19 | | | | | | |
| No | *Reference* | | *Reference* | | *Reference* | |
| Maybe | 1.41 (1.23 – 1.62) | <0.001 | 1.44 (1.25 – 1.67) | <0.001 | 1.33 (1.16 – 1.53) | <0.001 |
| Yes | 1.20 (0.99 – 1.46) | 0.063 | 1.46 (1.18 – 1.79) | <0.001 | 1.09 (0.90 – 1.33) | 0.372 |
| Diagnosed with COVID-19 | | | | | | |
| No | *Reference* | | *Reference* | | *Reference* | |
| Yes, and I still have it | 1.36 (0.64 – 2.90) | 0.415 | 1.84 (0.85 – 4.01) | 0.120 | 1.45 (0.69 – 3.15) | 0.333 |
| Yes, but I recovered | 0.88 (0.55 – 1.41) | 0.605 | 0.98 (0.59 – 1.60) | 0.938 | 0.87 (0.55 – 1.40) | 0.566 |

**Table B. Prevalence of specific worries for the overall sample and by pregnancy stage.**

| Worries | Pregnant | Postpartum | Overall |
|---|---|---|---|
| That my family members/friends will be infected with COVID-19 | 67.4% | 69.8% | 67.8% |
| That my family will not be able to visit me and the baby after delivery because of measures to prevent COVID-19 spread | 62.8% | 41.7% | 59.2% |
| That my partner will get COVID-19 and bring the infection home | 59.0% | 57.6% | 58.8% |
| That my unborn baby will get COVID-19 | 65.2% | 26.3% | 58.5% |
| That my partner/support person will not be able to be with me during delivery because of COVID-19 | 61.8% | 24.5% | 55.4% |
| That I will get COVID-19 and bring the infection home | 53.6% | 58.0% | 54.4% |
| That my parents/grandparents will not be able to visit the baby because of measures to stop COVID-19 | 51.4% | 49.2% | 51.0% |
| That COVID-19 will mean changes to my delivery plan | 45.4% | 17.9% | 40.6% |
| That the COVID-19 pandemic will significantly affect my economic situation/finances (for example, lose my job) | 38.3% | 36.0% | 37.9% |
| That I will not be able to have a baby shower or other baby celebration with family or friends | 36.2% | 26.6% | 34.5% |
| That my other children will get COVID-19 | 29.0% | 35.0% | 30.0% |
| Missing/canceling doctor's appointments | 26.5% | 25.9% | 26.4% |
| That I will not be able to breastfeed because of COVID-19 | 22.7% | 16.7% | 21.7% |
| That I will not be able to provide adequate childcare for my other kids | 18.8% | 23.1% | 19.6% |
| That I will not be able to attend the funeral of a family member | 19.3% | 18.9% | 19.2% |

**Table C. Prevalence of COVID-19 prevention behaviors for the overall sample and by pregnancy stage.**

| COVID-19 Behavior | Pregnant | Postpartum | Total |
|---|---|---|---|
| Washed your hands with soap or used hand sanitizer several times per day | 93.4% | 92.5% | 93.3% |
| Wore a face mask | 84.9% | 82.5% | 84.5% |
| Avoided public spaces, gatherings, or crowds | 82.1% | 85.2% | 82.6% |
| Avoided eating at restaurants | 69.3% | 74.2% | 70.1% |
| Avoided contact with people who could be high-risk | 66.1% | 68.4% | 66.5% |
| Disinfected surfaces around you | 64.6% | 60.6% | 63.9% |
| Canceled or postponed personal or social activities | 55.2% | 54.1% | 55.0% |
| Worked or studied at home | 50.8% | 40.3% | 49.0% |
| Prayed | 36.2% | 32.7% | 35.6% |
| Canceled or postponed work or school activities | 33.1% | 29.6% | 32.5% |
| Visited a doctor | 29.3% | 21.3% | 27.9% |
| Stockpiled food or water | 21.6% | 24.5% | 22.1% |
| Stockpiled hand sanitizer or disinfectant wipes | 19.1% | 25.7% | 20.2% |
| Canceled or postponed air travel for pleasure | 16.2% | 14.2% | 15.9% |

| | | | |
|---|---|---|---|
| Canceled a doctor's appointment | 11.9% | 16.2% | 12.7% |
| Canceled or postponed air travel for work | 8.0% | 9.6% | 8.3% |
| Stockpiled medication | 7.1% | 9.6% | 7.5% |

**Supplementary Materials (S2)**

**COVID-19 Worries**

Exploratory factor analysis using the tetrachoric correlation matrix of the COVID-19 worries questionnaire found four meaningful factors **(Fig A, Fig B, Table A)**. The Kaiser-Meyer-Olkin (KMO) measure of 0.80 indicated a moderate sampling adequacy. Both the scree plot and parallel analysis suggested 4 underlying factors. The first factor was characterized by high loadings of worries associated with social factors (e.g., parents/grandparents unable to visit, family unable to visit, not able to have a baby shower/other baby celebration with family and friends). The second factor encompassed worries related to COVID-19 infection (e.g., participant bringing infection home, partner getting infected and bringing infection home, family/friends getting infected with COVID-19). The third factor included child-related items (e.g., not able to provide adequate childcare for other kids, other children may get COVID-19, not able to breastfeed). Finally, the fourth factor represented delivery-related worries (e.g., partner not present during delivery, changes to delivery plan, not able to breastfeed, unborn baby will get COVID-19). Two items that did not load onto any of the factors (missing doctor appointments, economic/finances) were examined individually. As such, a total of six clusters of worries were examined in subsequent analyses.

**Fig A. Tetrachoric correlation matrix of the COVID-19 worries questionnaire.**

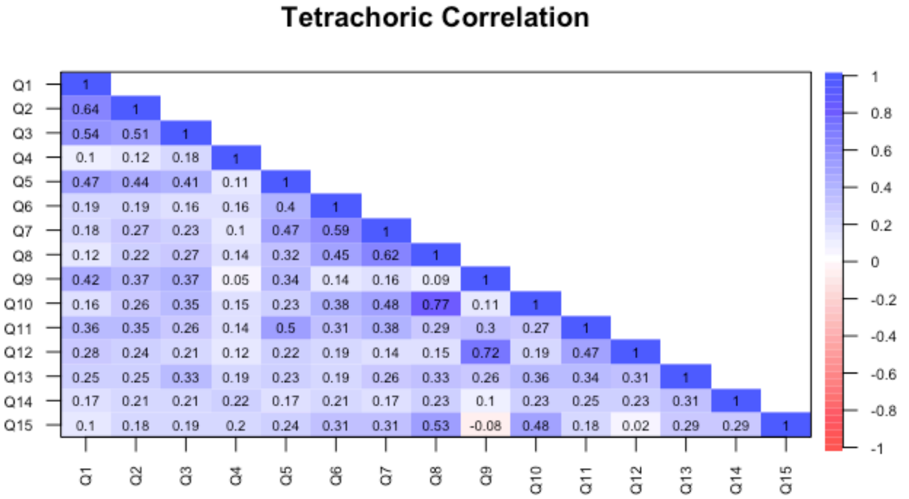


Q1 – participant will bring infection home; Q2 – partner will bring infection home; Q3 – family/friends will be infected with COVID-19; Q4 – COVID-19 will significantly affect economic situation/finances; Q5 – unborn baby will get COVID-19; Q6 – COVID-19 will cause changes to delivery plan; Q7 – partner will not be present during delivery because of COVID-19; Q8 – family unable to visit; Q9 – other children will get COVID-19; Q10 – parents/grandparents unable to visit; Q11 – not able to breastfeed because of COVID-19; Q12 – not able to provide adequate childcare for other kids; Q13 – not able to attend the funeral of a family member; Q14 – missing doctor appointments; Q15 – not able to have a baby shower/other baby celebration with family and friends

**Fig B. Scree plot and parallel analysis of the COVID-19 worries questionnaire.**

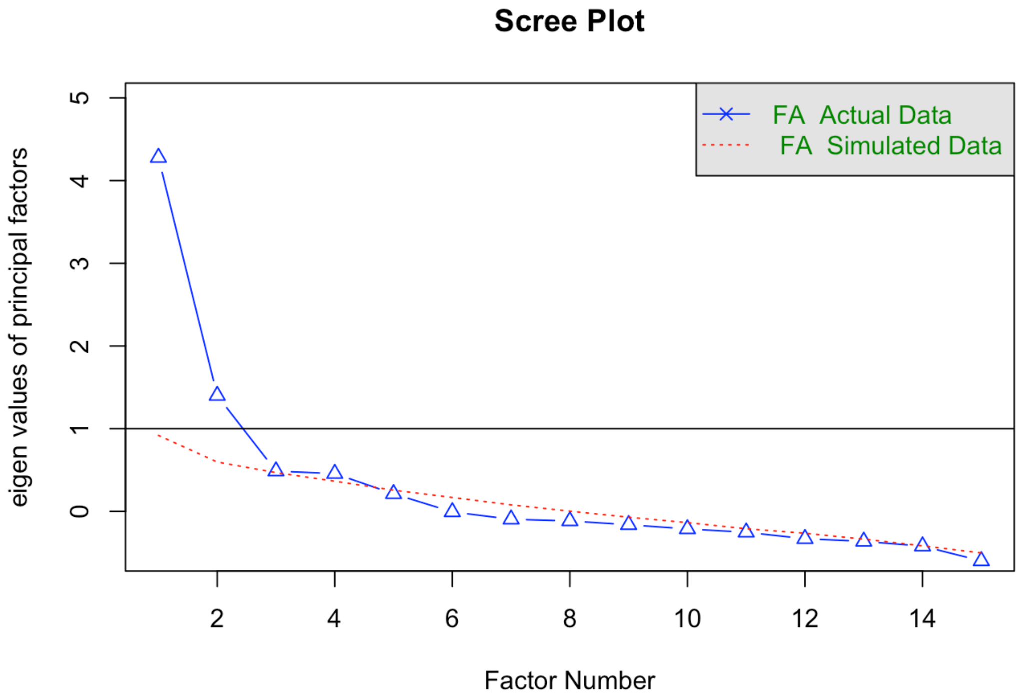


**Table A. Factor loadings[a] based on a tetrachoric factor analysis with oblimin rotation of the COVID-19 worries questionnaire.**

| Worried that | Factor 1: Social | Factor 2: Infection | Factor 3: Child | Factor 4: Delivery |
|---|---|---|---|---|
| Parents/grandparents unable to visit | 0.88 | | | |
| Family unable to visit | 0.78 | | | |
| Not able to have a baby shower/other baby celebration with family and friends | 0.53 | | | |
| Not able to attend the funeral of a family member | 0.33 | | | |
| Missing doctor appointments | | | | |
| COVID-19 will significantly affect economic situation/finances | | | | |
| Participant bring infection home | | 0.80 | | |
| Partner will get COVID-19 and bring the infection home | | 0.71 | | |
| Family/friends will be infected with COVID-19 | | 0.68 | | |
| Not able to provide adequate childcare for other kids | | | 1.02 | |

| | | | |
|---|---|---|---|
| Other children will get COVID-19 | | 0.66 | |
| Partner will not be present during delivery because of COVID-19 | | | 0.72 |
| COVID-19 will cause changes to delivery plan | | | 0.62 |
| Unborn baby will get COVID-19 | 0.41 | | 0.56 |
| Not able to breastfeed because of COVID-19 | | 0.35 | 0.37 |

[a]All loadings >.30 appear in the table
